# Cross-sectional equity analysis of accessibility by automobile to tertiary care emergency services in Cali, Colombia in 2020

**DOI:** 10.1101/2022.02.03.22269929

**Authors:** Luis Gabriel Cuervo, Eliana Martinez Herrera, Lyda Osorio, Janet Hatcher Roberts, Daniel Cuervo, María Olga Bula, Luis Fernando Pinilla, Felipe Piquero, Ciro Jaramillo, the AMORE Project Collaboration

## Abstract

This study provides data on equity in accessibility to tertiary care emergency services in Cali, accounting for traffic congestion, in two separate weeks in 2020.

This cross-sectional study builds on a proof-of-concept, the [AMORE Project](https://osf.io/4esnz/)^(1)^ and provides a baseline assessment of accessibility to urgent tertiary care at peak and free flow traffic times in Cali.^1^ It makes the case for assessing travel time over distance, and accounting for traffic congestion.

This study indicates that people in vulnerable situations have to travel longer and therefore invest more of their personal direct and indirect resources to access tertiary care emergency departments than the average population. This study emphasizes the added value of integrating new data sources that can inform health services and urban planning. These new data sources merit future testing by concerned stakeholders.^1^

This study used the digital AMORE Platform to show the effects of traffic congestion on equitable access to tertiary care emergency departments. The data shows which populations take longer to reach a facility within a time threshold under different traffic congestion levels.

The broader proof-of-concept assesses the value of new data obtained by integrating secondary data from publicly available sources. These sources combine geospatial analysis with census microdata, health services location data, and bigdata for travel times.

The analysis covered the city of Cali, which has 2.258 million residents and is the third-largest city in Colombia. The analysis shows the projected accessibility assessments for two weeks during the COVID-19 pandemic, 6 – 12 July 2020, and 23 – 29 November 2020. Restrictions on car travel had been lifted before the July assessment, but stay-at-home orders were in place during the November assessment, which showed substantially less traffic.

This assessment found that traffic congestion sharply reduces accessibility to tertiary emergency care. Reduced access has the greatest impact on people with less education, those living in low-income households or on the periphery of Cali, and specific ethnic groups (e.g., nomadic people like the Rrom, and Afro-descendants). This assessment also identifies the concentration of tertiary care emergency departments in areas of lower population density, leaving large swaths of the population with poor accessibility.

Data was reported in dashboards that used simple univariate and bivariate analyses. In July 2020, the estimated overall accessibility at peak traffic hours was 37% and in November 2020 it increased to 57% due to reduced traffic congestion. These results illustrate the value of the proposed tools in monitoring and adjusting to changing conditions.

## Introduction

### Background/rationale

Every minute counts in life-threatening emergencies (e.g., important blood vessel obstructions or ruptures, airway obstructions, asphyxiation or drowning, severe trauma, serious wounds) that do not leave time for referrals. The wellbeing of patients depends on getting immediate attention in a tertiary care facility. These facilities provide subspecialized care and access to highly trained personnel, sophisticated surgical theatres and intensive care.

This study delivers a baseline assessment of accessibility by automobile to such services in Cali, Colombia. This study assessed traffic congestion for Cali residents traveling by automobile (private or for-hire), which is how residents typically reach tertiary care facilities in emergencies.

Accessibility is a dynamic spatial attribute measured as the travel time needed to reach a health service from the origin of the demand.^2–7^ It is dynamic because it changes as travel times fluctuate with traffic congestion. Poor accessibility is a recognized barrier to health equity that has until now been difficult to study and monitor.^5,8^

Traditional assessments of accessibility in urban planning seldom consider that accessibility is dynamic. Origin-destination studies and surveys lack a dynamic assessment of how traffic congestion or changes in infrastructure or populations affect accessibility.^9,10^ These traditional assessments are usually done every five to ten years and are expensive. The conditions they assess may have changed by the time results become public, which can render any proposed solutions irrelevant.^11,12^ This study integrated new data and exposes the important links between equity and accessibility to tertiary care emergency services.

These innovative approaches for integrating data and accessible web-based platforms offer an important opportunity for evidence-based decisions and planning to improve health coverage. These approaches capitalize on, for example, big data from smartphones, which can feed accurate travel time estimates. We could therefore use dynamic, affordable, and updatable assessments that account for traffic congestion, thus focusing on travel time to hospitals instead of distance from them.^6,10,12–16^ We also integrated equity-relevant data that we used to perform equity analyses.

### Objectives

This study aims to characterize accessibility to tertiary care emergency health services in urban Cali and the relationship of accessibility to sociodemographic factors relevant to health equity.

## Methods

### Study Population and setting

This study assesses an aspect of emergency care: emergencies requiring attention in tertiary care institutions. By early July 2020, COVID-19 pandemic-related quarantine and stay-at-home orders had been lifted and traffic projections showed substantial congestion. By November 2020, these measures had been reinstated, car travel was restricted by license number, and traffic projections showed a reduction in travel times.^17–19^

This cross-sectional study was carried out using data downloads obtained in the urban area of the city of Cali, in Southwest Colombia. Cali is the third-largest city in Colombia and the largest urban center in the country’s southwest and Pacific regions, with 2.258 million residents in 2019. About half of the population lives in low-income households, 42% in middle-income households, and 8% in high-income households. Housing stratification does not necessarily represent the income of residents. For example, domestic workers living in mansions and receiving the minimum wage would still be counted as living in a high-income stratum.^20,21^

About 84% of the population identifies as white, 14% as Afro-Colombian, and a small proportion as indigenous or nomadic people like the Rrom. In December 2020, unemployment rates in Cali were 26.7% for women and 18.5% for men, a one-year increase of 12.5% and 8.8%, respectively. The situation is worse among young people and an estimated 52% of women and 47.2% of men rely on working in the informal economy. The COVID-19 pandemic punished the local economy. While 1 in 5 people are unemployed, the rates are higher among people in low-income households. From 2016 to 2020, Cali also absorbed 139,000 migrants from Venezuela, 25,000 in 2020 alone.^22^

Poverty, inequity, and discrimination drove social unrest that led to violence after a 2021 national strike.^23,24^

The city government is dividing its 22 communes into a six to eight districts, which might lead to negotiations over resources and issues such as access to essential services. ^25,26^

Looking to the future, identifying and proposing public policy plans and partnerships could improve health equity and bring hope to residents. These measures could also reduce social injustices, including the burden of the inverse care law that vulnerable populations pay more to access essential health services.^5,27,28^

### Targeted Sites/Participants

We targeted the 14 tertiary care institutions with emergency departments registered in the Ministry of Health Special Registry of Health Services Providers (REPS in Spanish). We searched the registry twice, in July 2020 and January 2021. We verified that all tertiary care institutions provide surgery and intensive care services and listed those institutions on Map 1.^29^

**Map 1.**
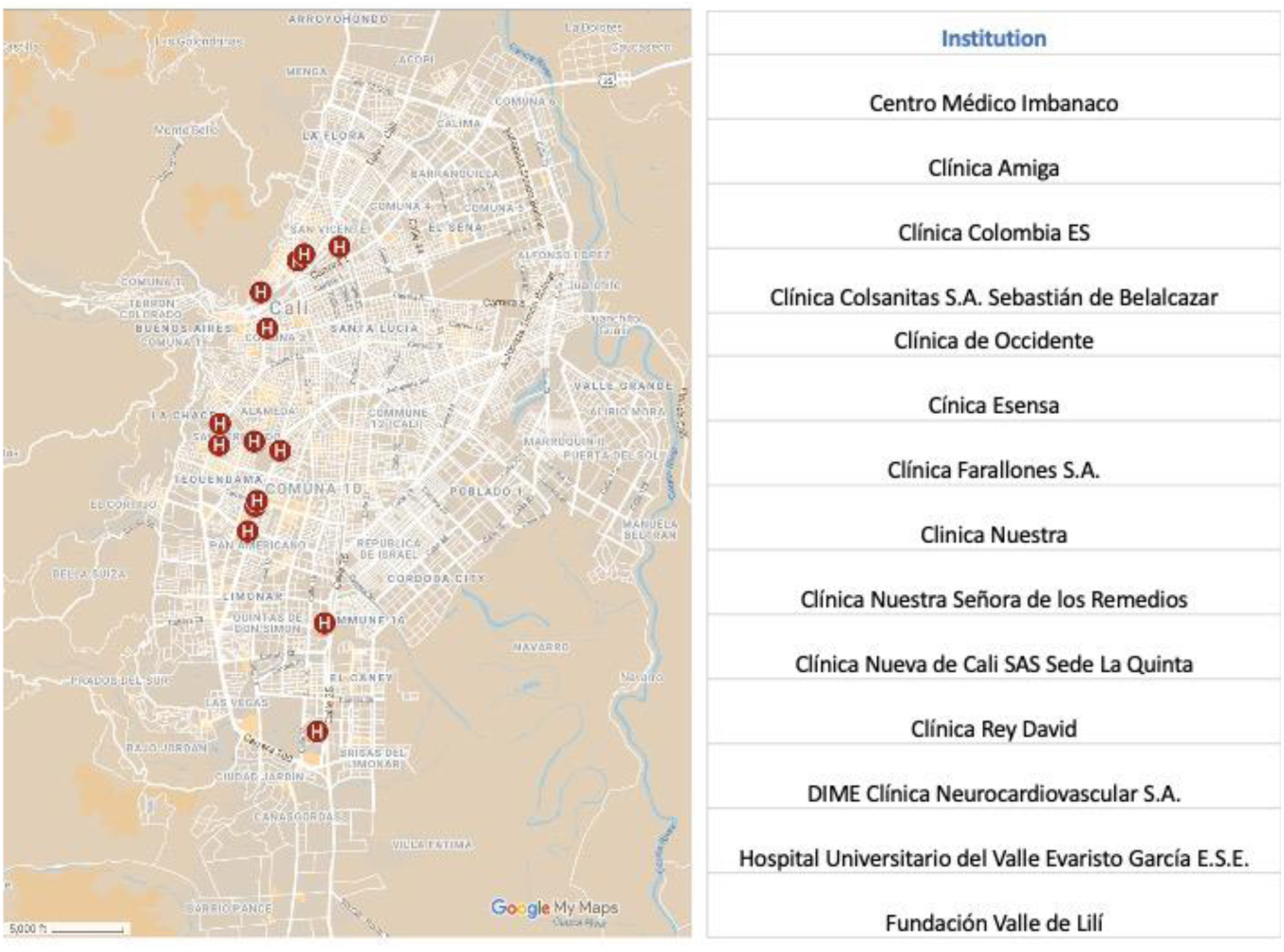
Location of tertiary care hospitals with an emergency department (order in REPS displayed them) ^29^

### Study design

This is a cross-sectional study that used digital technologies and analytics to integrate publicly available data sources. The study generated new knowledge that shows the potential value of examining an evidence-based approach to accessibility and health equity. The study used updatable data to assess travel times and evaluate the effects of interventions and changes in the infrastructure, service provision, traffic congestion, and population.

### Study methods

- Used dynamic assessments of travel times to account for traffic variations.
- Used input from diverse stakeholders to create an interactive platform that displays intersectoral data on dashboards so stakeholders can interpret data quickly and accurately.
- Offered disaggregated data to enable straightforward equity analysis of accessibility.
- Enabled situational analysis of accessibility in an urban setting and supported monitoring and evaluation of health equity related to accessibility.

The cross-sectional study data was obtained from an internet-based platform, the AMORE Platform (https://www.iquartil.net/proyectoAMORE), hosted by iQuartil SAS, an analytics company, and developed under the leadership of the principal investigator (LGC).

The AMORE Platform integrates data from:

- 2018 National Census Data for Cali, obtained from the public official databases of the Colombian National Department of Statistics – DANE.^20,21^
- The administrative divisions of Cali obtained from Colombia’s [IDESC Geoportal](https://idesc.cali.gov.co/geovisor.php), Traffic Analysis Zones (TAZ), and census block sectors.^11,30^
- Google’s Distance Matrix API. For this baseline assessment, of the urban area of Cali, we downloaded the data on July 3, for the week of 6 – 12 July 2021. For the week of 23 – 29 November 2020, the predicted times were downloaded on October 27, 2020. Travel times varied substantially during the COVID-19 pandemic and it is unclear how this influenced Google Distance Matrix algorithms.^31^ Empirical and anecdotal reports suggest they remained accurate.
- The 14 tertiary care institutions with emergency department in Cali, identified using REPS.^29^

Databases were integrated and tested between August 2020 and October 2021 using KNIME® and Python™ software (back end) and the interface (front end) was developed in Microsoft PowerBi™.

## Results

The AMORE Platform dashboards and visualizations provide simple indicators such as colored maps, dials, bars, and data. These indicators show travel time to the nearest tertiary care emergency room and descriptive statistics for each urban sector at a given traffic congestion level.

The AMORE Platform displays a situational analysis of accessibility in simple visualizations with filters that let users disaggregate data by sociodemographic characteristics for an equity analysis.^32^ The top of the Platform filters scenarios (i.e., tertiary care emergency departments) and census adjustments (Figure 1). It has nine traffic congestion levels represented with the schedules for a regular week. A dial shows the share of the population within travel time thresholds set in a slider.

**Figure 1.**
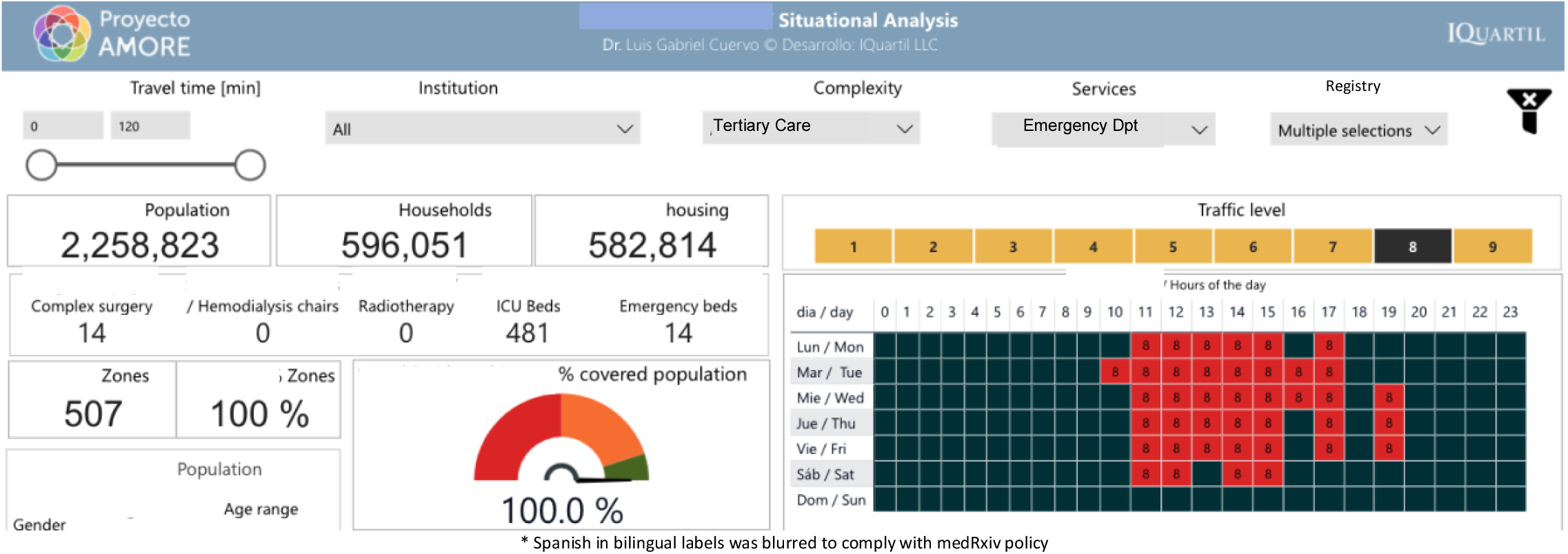
Top section of the AMORE Platform situational analysis

The middle section displays a population pyramid and maps with the 14 tertiary care emergency departments, travel times, and population density (Figure 2). The choropleth maps can be expanded and rotated for 3D-display that uses the height of sectors to represent population density (Map 2).

**Figure 2.**
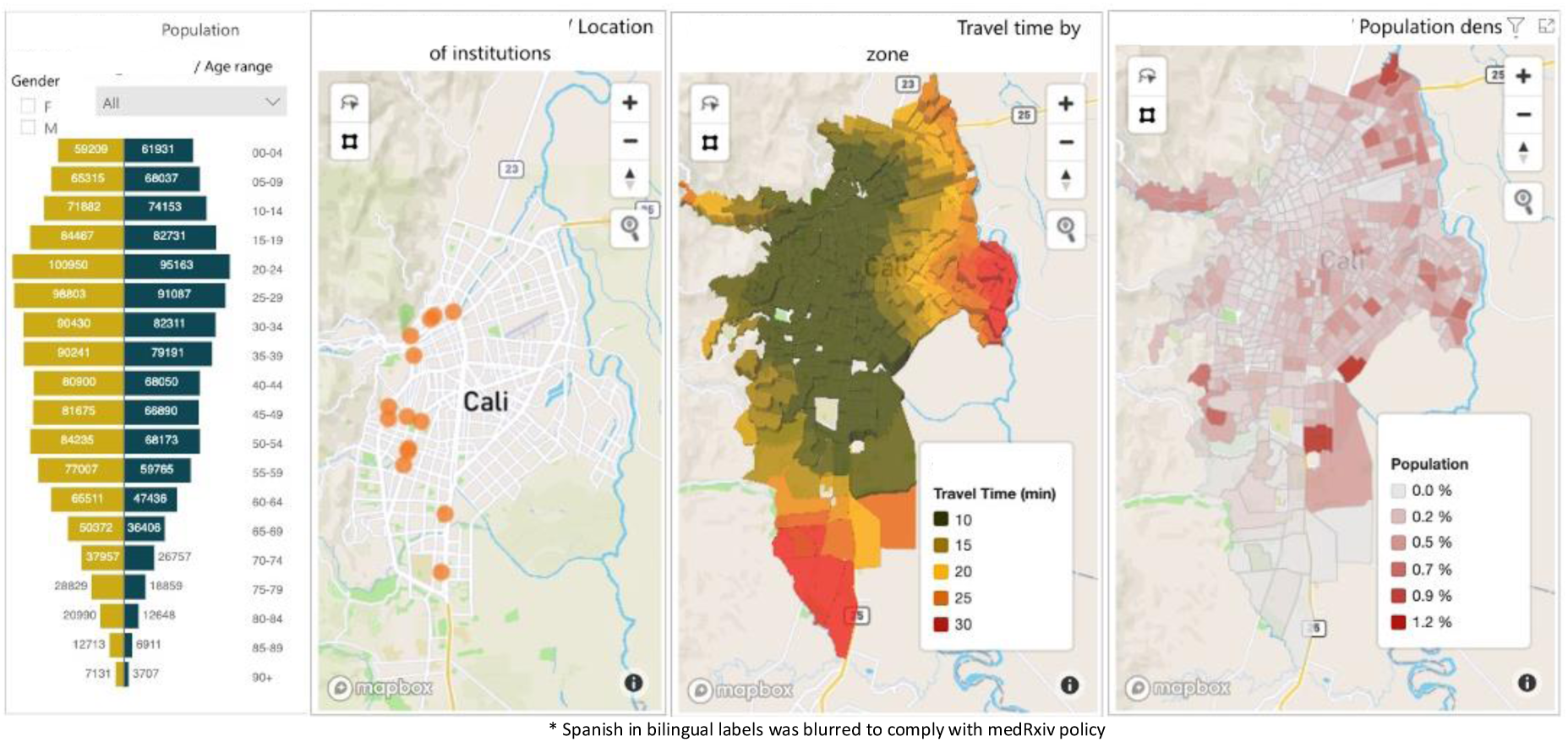
Mid-section, Situational analysis of the AMORE Platform

**Map 2.**
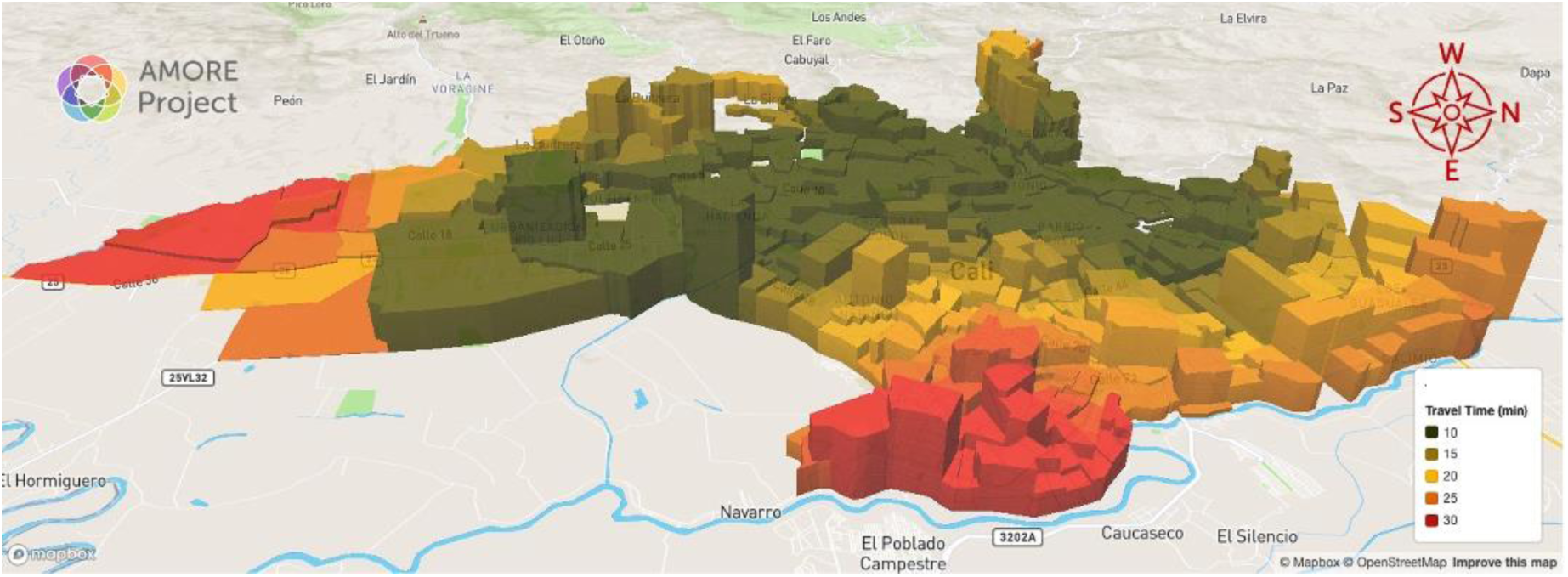
Travel times and population density, tertiary care institutions, peak hour, November 2020

The choropleth maps consist of 508 TAZs established by MetroCali (Integrated Massive Transit System) in 2015 for the urban area and linked to the geotagged census block information, matching the population with these TAZs.^11^ The origin-destination times were estimated from the population-adjusted geographic centroid of each TAZ to the centroid where each institution was located.

In 2019, Colombia’s National Department of Statistics recommended adjusting the 2018 population of Cali upward by 18% from the original census data because of underregistration.^21,33^ To make the adjustment, a random selection of 18% of the individual records from the unadjusted census were duplicated with all their original information, adding 495,219 people to complete the 2,258,823 records in the adjusted census. In verifying the records, we found a matching distribution of the variables and results with the unadjusted census. The AMORE Platform lets users toggle the census adjustment (Figure 1, “Tipo de Registro”).

The bottom section displays sociodemographic characteristics. The graph bars activate filters for data on selected demographics (Figure 3).

**Figure 3.**
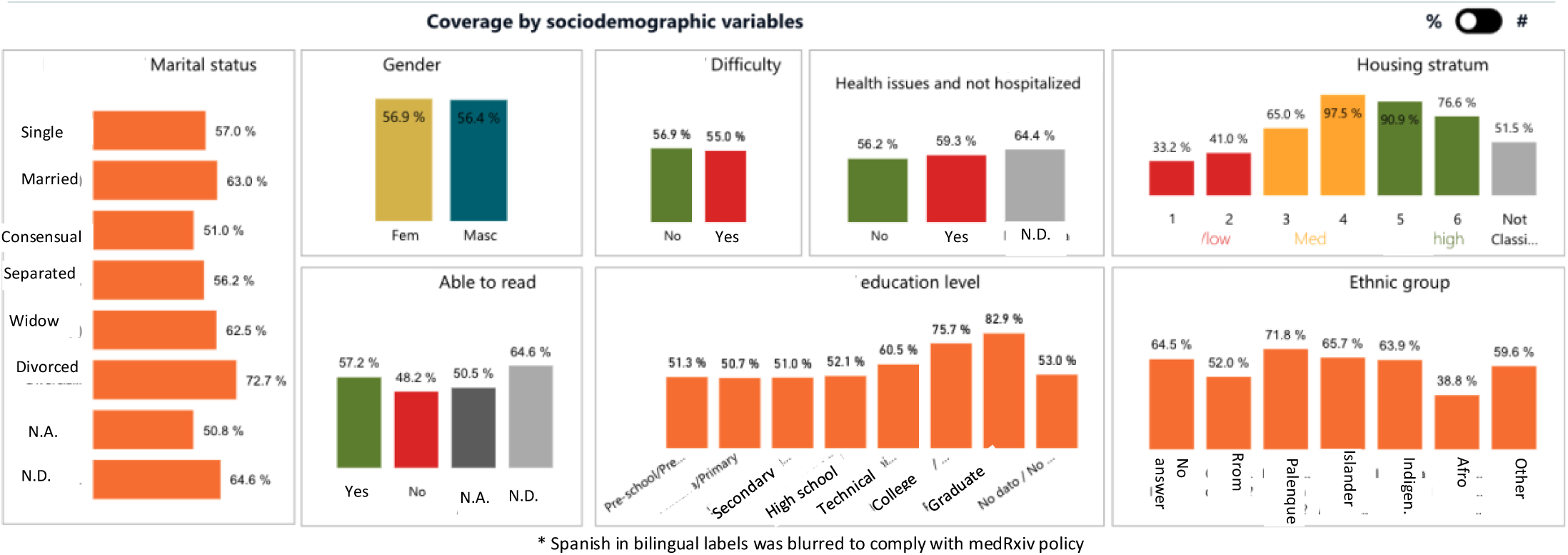
Demographic characteristics of the populations that can reach a tertiary care hospital within 15 minutes, Nov. 2020.

### Variables

The AMORE Platform displays the absolute and relative figures for the georeferenced data. This is done for the city or for selected TAZs within a travel time schedule (Figure 4). The variables integrated in the platform are listed in Table 1.

**Figure 4.**
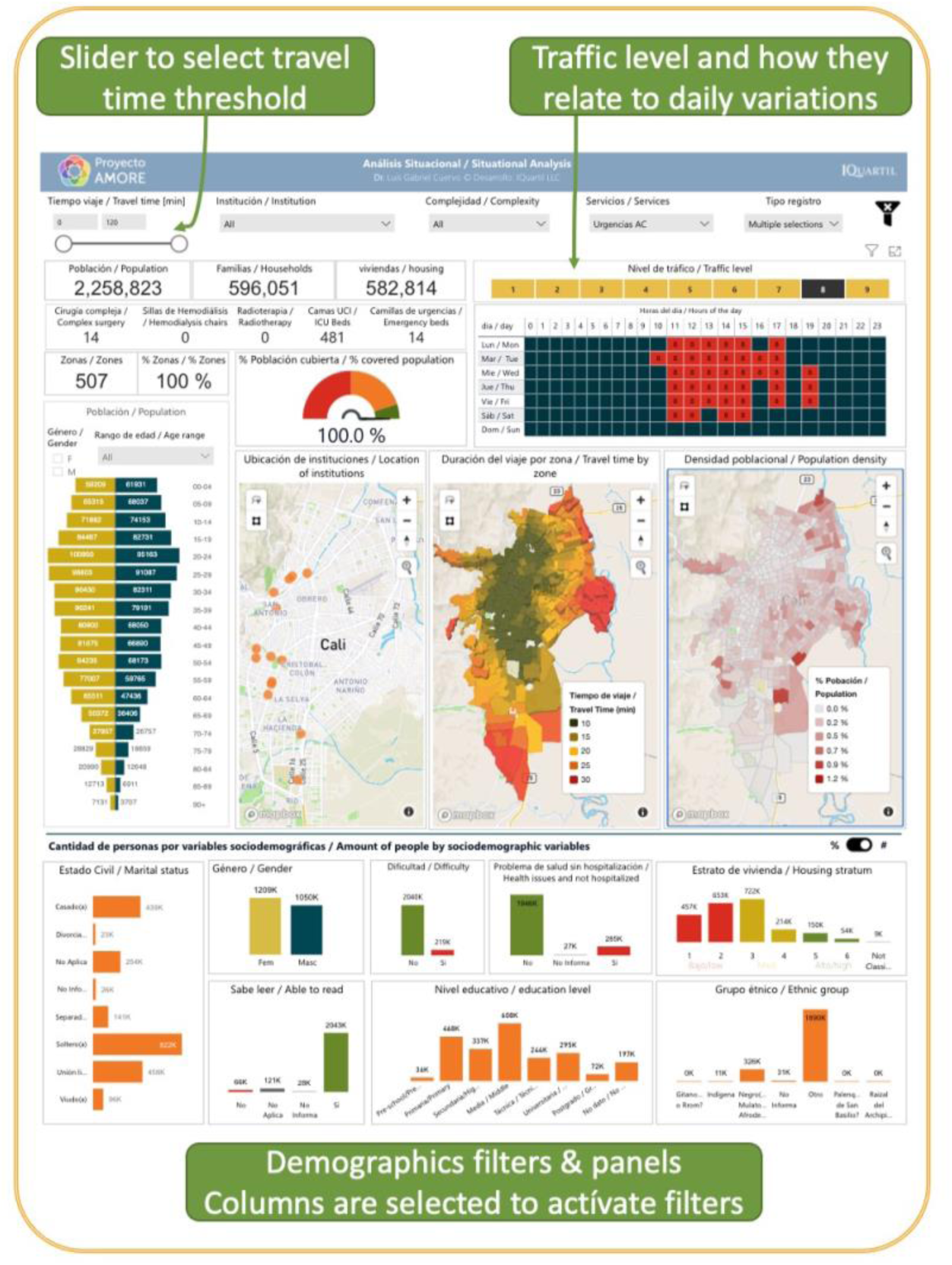
Situational analysis in the AMORE Platform, filters, visualizations (data July 2020)

**Table 1.** Census data included in the AMORE Platform dashboards and maps

| Geotagged variables | Platform display |
| --- | --- |
| Age in completed years grouped by quinquennium (census) | Population density per Sector/TAZ |
| Ethnicity, self-described | Health service: tertiary care, radiotherapy, hemodialysis |
| Health status (Sick / Healthy) | Optimized location if expanding 1-3 health facilities |
| Highest education level attained | Absolute and relative figures of modified aggregation |
| Literacy | Travel time thresholds (slider + choropleth heatmap) |
| Marital status | Travel time thresholds (slider + choropleth heatmap) |
| Population pyramid by gender and age | Travel times |
| Report of disability / physical condition | Absolute and relative figures |
| School attendance | Absolute and relative figures |
| Household inhabitants | Average for the selected population |
| Housing inhabitants | Average for the selected population |
| Housing socio-economic stratum | Absolute and relative figures for selected population |
| Housing type | Absolute and relative figures for selected population |

The census was done by interviewing an adult in each household. Data was stored linking it to a city block code to anonymize it. The AMORE Platform used the census microdata categorizations and, for a few variables, aggregate groups for simplification (e.g., education was simplified with guidance from an expert in Colombia’s education system, Psychologist Myriam Lorena Rosero Hernández, ME).

### Data sources / measurements

We used the controls listed in Table 2 to conduct univariate and bivariate analyses.

**Table 2.** AMORE Platform displays resulting from integrating travel times, services, origin sectors, and census data

|  |
| --- |
| **Variables that change according to traffic, travel time threshold, and other filters** |
| Filter and slider to select travel time threshold for the analysis. |
| Drop-down list with institutions that can be toggled for inactivation. |
| Drop-down list to select people registered by the census, the 18% adjustment, or the total adjusted census data. |
| Traffic levels according to clusters identified with a K-means clustering algorithm. |
| Indicator of % of the total of the population covered per selected thresholds, traffic level, and other filters. |
| Absolute and relative figures provided for each variable. |
| Maps are organized according to TAZs or sectors. |
| Intensive care beds data was taken from REPS for each hospital and included in the database. |
| **People with accessibility for a selected time threshold and traffic level.** |
| **Blocks with accessibility for a selected time threshold.** |
| **TAZs within the time threshold by level of traffic.** |
| Household inhabitants |
| Housing inhabitants |
| Housing by power bill economic stratum |
| Housing type |

### Bias

Each source is susceptible to biases and imprecisions, but these are unlikely to change our conclusions substantially. Some of the data sources and the timing of their updates can introduce a source of bias. For example, the census is updated every five years, and there has been a significant flow of migrants to Cali since the last census, in 2018, and job losses rose during the COVID-19 pandemic.^22,34^ These developments likely resulted in some internal displacement and may make our results more positive than reality.

Traffic patterns may have changed with the imposition and lifting of pandemic-related restrictions, thus altering traffic predictions. Stay-at-home orders and traffic restrictions may have reduced traffic congestion, causing an overestimate of accessibility. Google Distance Matrix API may have more accurate travel times for areas where more people travel with mobile phones.

Populations are not evenly distributed across TAZs. We therefore adjusted TAZ geographic centroids by weighing the population distribution. Because centroids had irregular forms, population weighed centroids could end outside the boundaries of a TAZ. This required relocating the adjusted centroid to the nearest border, generating some imprecision that could likely result in some seconds or minutes of imprecision in travel time estimates.

It is possible that the relevance of our findings could change if a new tertiary care facility is registered in REPS (i.e., a new institution opens, or an existing institution is reclassified as tertiary, changing the results). However, the interactive platform would allow for prompt updates and reanalysis in response to these contingencies.

Income categorization is determined by the individual household electricity bill, which is graded from 1 to 6, with 6 representing the highest-income households. It is possible that some households were misclassified (e.g., due to error or corruption) and that low-income people are living in higher-income households. Low-income people may live with relatives or work as maids or support staff. This kind of misclassification could introduce some bias by representing low-income populations as having higher income.

We developed nine traffic clusters for the city. Traffic patterns are not homogeneous within a city or sector; traffic flow patterns vary in time and direction. The nine traffic congestion levels for Cali were sorted in incremental order for each TAZ so that level 9 would always represent the heaviest traffic and level 1 the lightest (Chart 1). This shows the typical times at which traffic is higher in the city, but it is not in every single sector of the city, as these have their own variations. For example, traffic congestion at noon on Saturdays and early evenings of weekdays affects almost every sector of Cali, but not all. Similarly, commuting at different times generates different directional patterns.

The Colombian census recognizes ethnicity, but some people likely found it difficult to choose their ethnicity. The census lacks an option for Colombians of white or mestizo descent, two large groups not specifically listed on the census. Similarly, people with multiethnic parents may find it difficult to choose one ethnic category.

The definition of an acceptable travel time threshold to reach a tertiary care emergency department is arbitrary. For this analysis, we chose a threshold of 15 minutes at peak traffic congestion times. Notably, the distribution of traffic levels is skewed towards heavy congestion from Monday to Saturday between 6:00 and 22:00, with a mode of traffic level 8 (40/168 hours in the week, 24% of the time)

**Chart 1.**
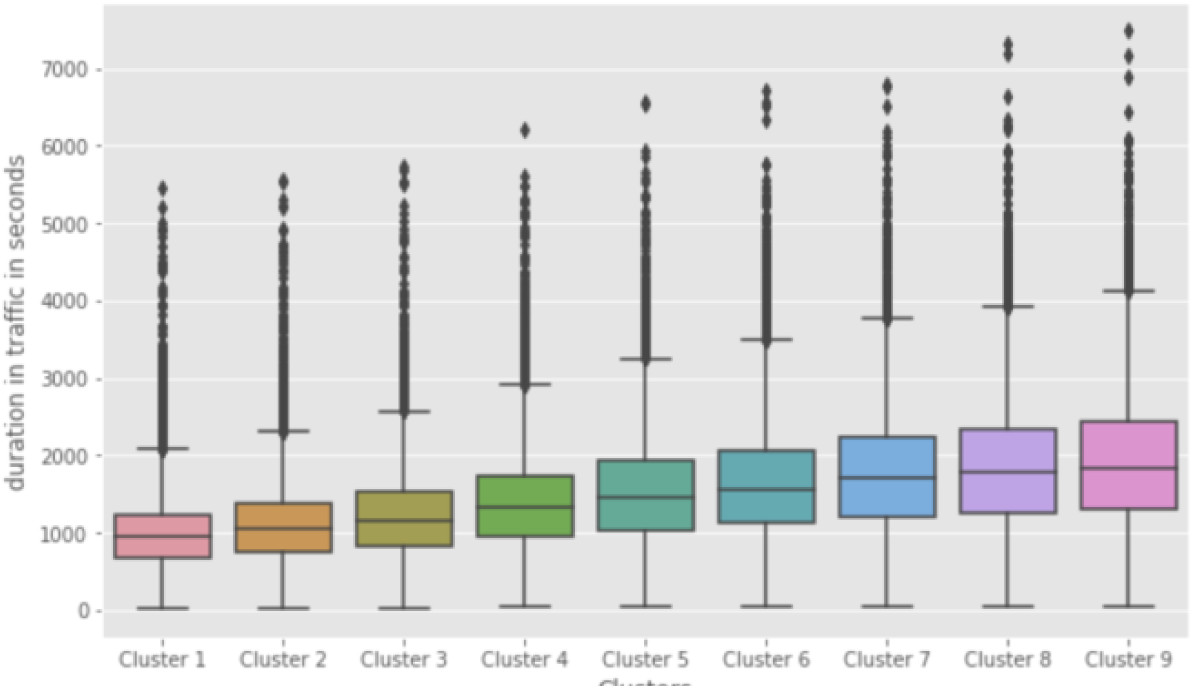
Travel-time clusters (Cluster 0 = 1 in the platform, Cluster 8 = 9 in the platform)

**Chart 2.**
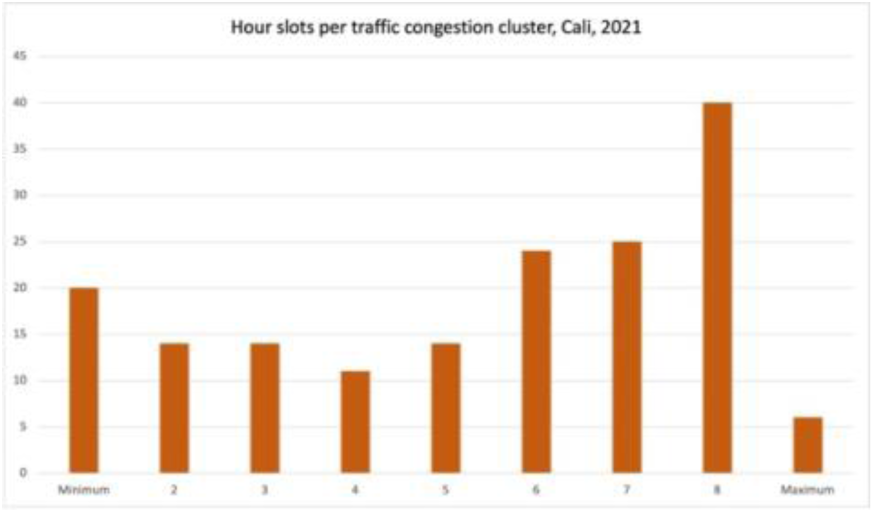
Hours of the week per cluster

The 168 hours in a week are distributed in the 9 clusters in Chart 2, showing that heavy traffic is the norm in Cali.

## Results / Outcomes

### Participants

The study included all the population data from the adjusted, 2,258,823 people from 596,051 households, living in 582,814 housing units. Most of the population is mestizo or white (83.7 %) or Afro-descendants (326,492; 14.5%). Islanders and Rrom people represent less than 1% of the population.

### Descriptive data

The analysis found that with traffic, most of the low-income population was unable to reach the nearest tertiary care emergency department within 15 minutes, whether in November 2020 or July 2020 (Map 2 and Map 3, respectively). The analysis also shows how accessibility is an access barrier for people living in low-income households, in areas with high population density, and for those living in the peripheral sectors.

**Map 3.**
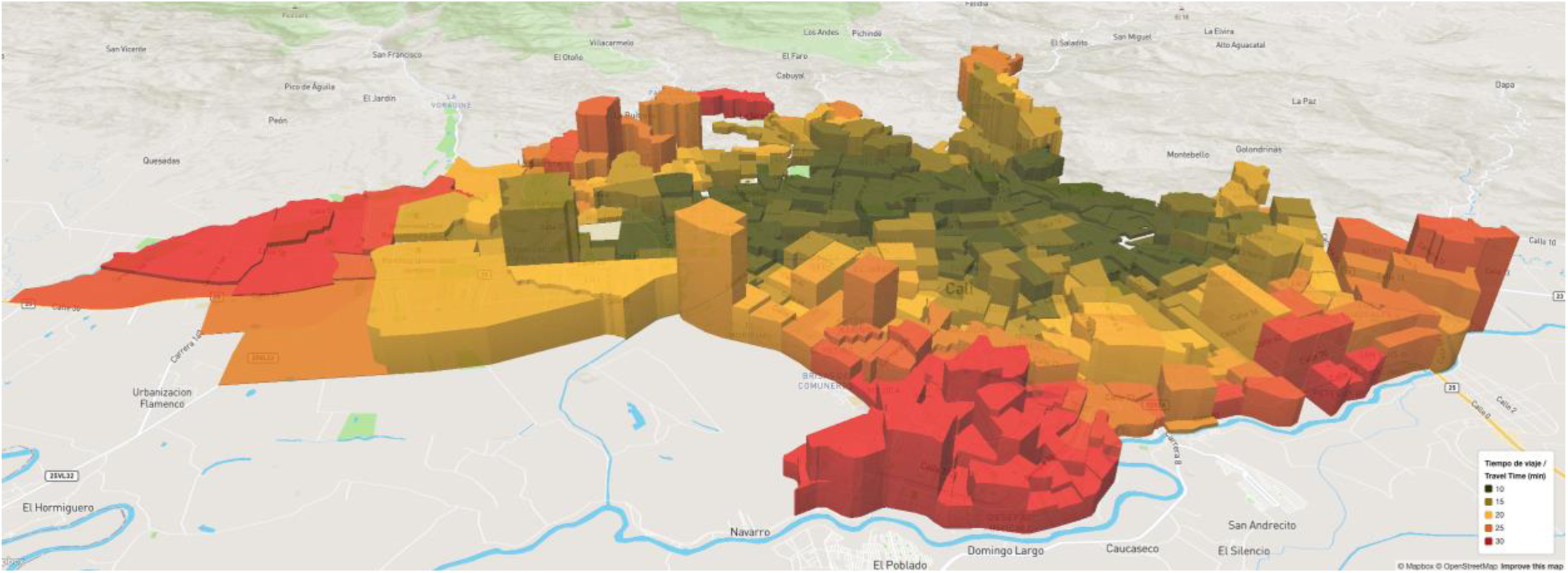
travel times and population density (height). Orientation: West top, North right. Source: AMORE Platform, Situational Analysis July 2020

### Main results

The effects of traffic disaggregated by household income level, ethnicity, gender and age, education level, and civil status are presented in Chart 3 and displayed in Figure 4, Figure 5, and Figure 6.

**Figure 5.**
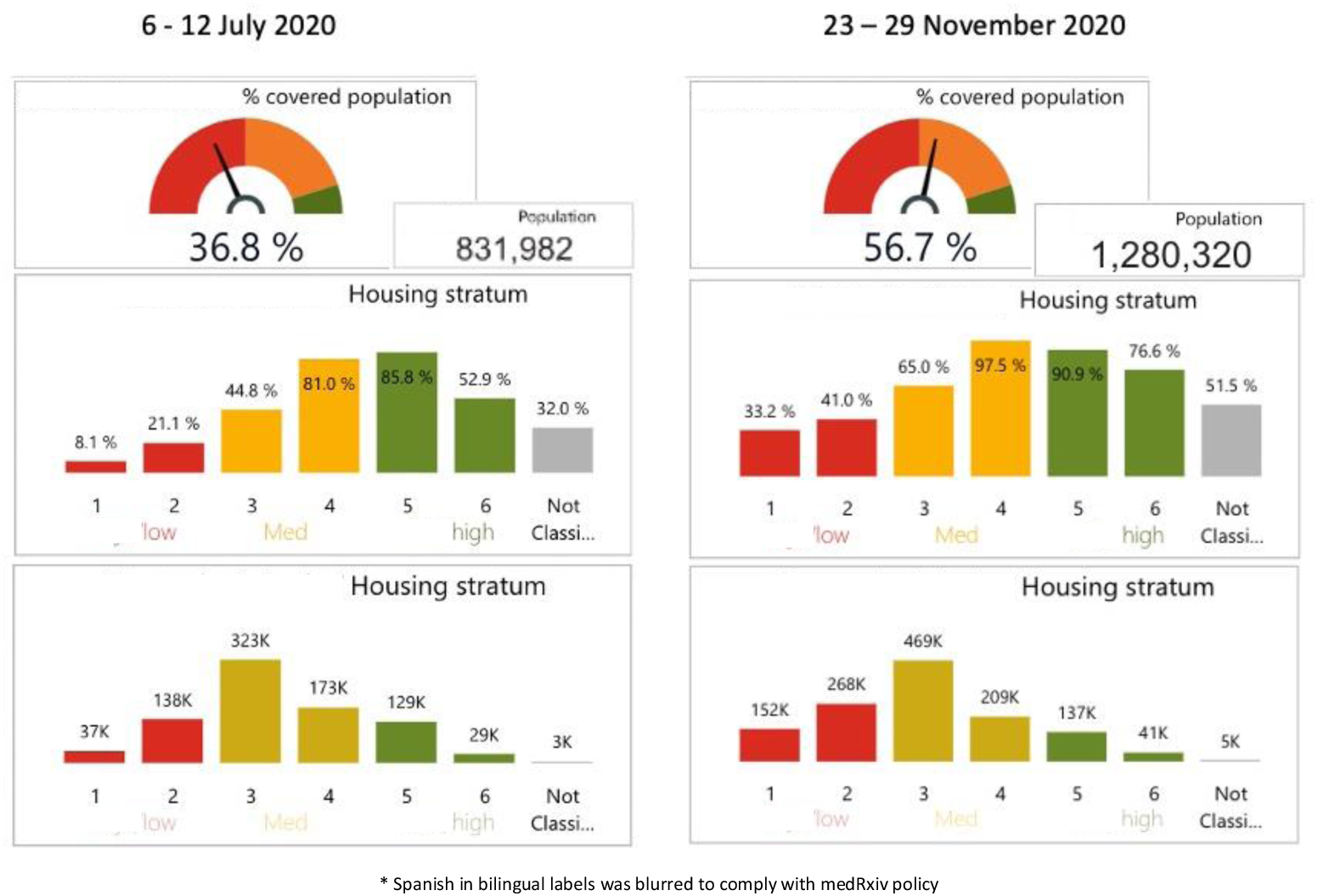
Changes in 15’ accessibility to tertiary care institutions by income; July and November 2020

**Figure 6.**
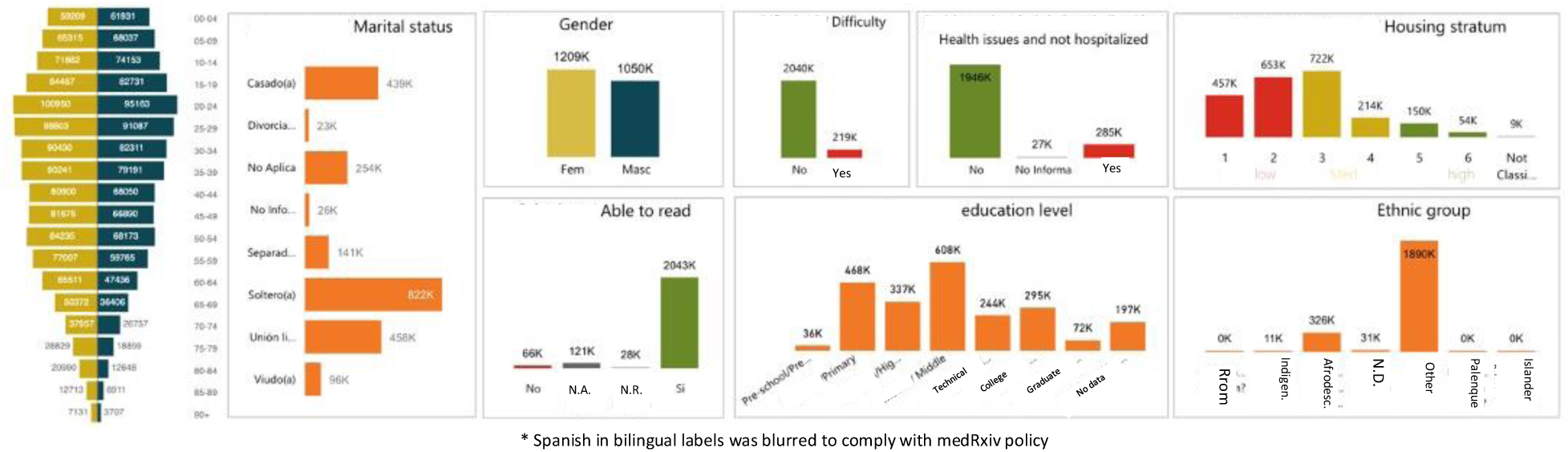
Sociodemographic characteristics of the population of Cali (2018 adjusted census data) absolute figures

### Traffic variations and their effect (July vs November 2020)

#### 6 – 12 July vs 23 – 29 November 2020

While the July travel time predictions pointed to 831,982 people (36.8%) living within 15 minutes of travel time from tertiary care emergency services, in November this increased to 1.28 million (56.7%). The distribution of the absolute and relative figures when disaggregating data by income level indicated lower accessibility for the poor and those living in peripheral sectors (Figure 5 and Chart 3, and evident in Map 2 and Map 3 to those familiar with the demographic distribution of Cali). These populations also have a higher representation of people from minority ethnic groups and people with lower educational attainment.

The characteristics of the census population are shown in Figure 6. The variable “Difficulty” represents self-assessed disability; the ethnic group “otro” [other] included Caucasian descent and mestizo populations.

Chart 3 Shows the data obtained from the platform for the July 2020 and November 2020 assessments, which lets users explore equity considerations.

### Other analyses

Figure 8 compares accessibility by socio-economic stratum at peak and free flow traffic congestion. It illustrates how people living in low-income households have longer travel times and are more impacted by traffic congestion, forcing them to invest more resources in accessing services.

**Figure 7.**
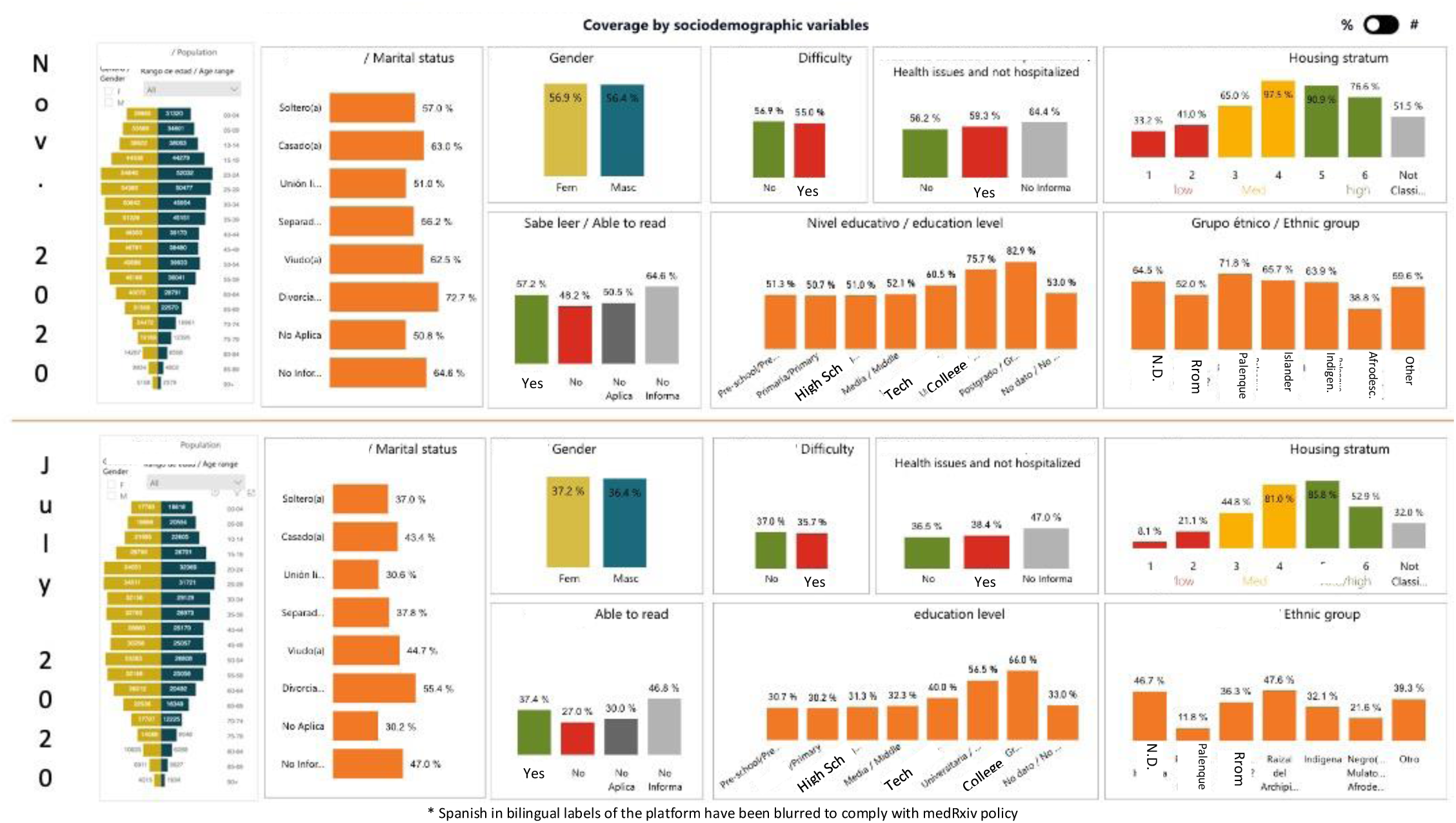
Characteristics of the population within 15 min of a tertiary hospital Jun vs. Nov 2020, relative figures

**Figure 8.**
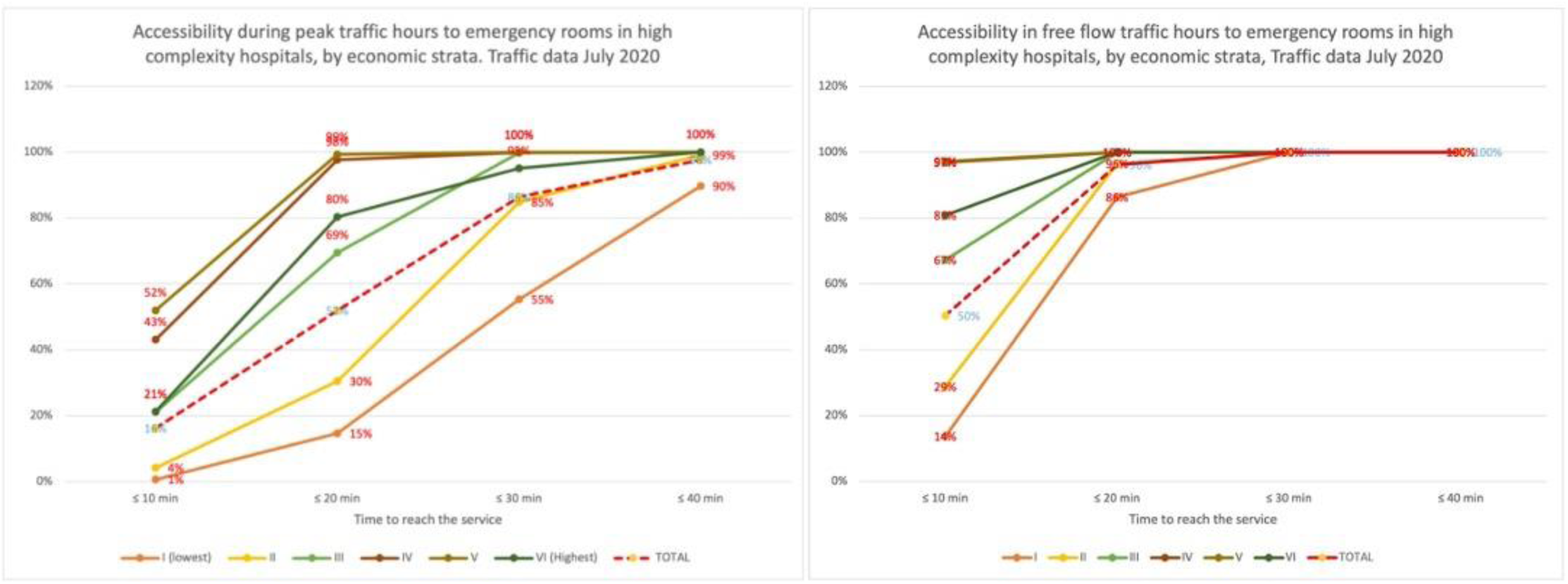
Accessibility peak vs. free flow hours, by housing stratum (I-II are low income, V-VI high income), July 2020

**Chart 3.**
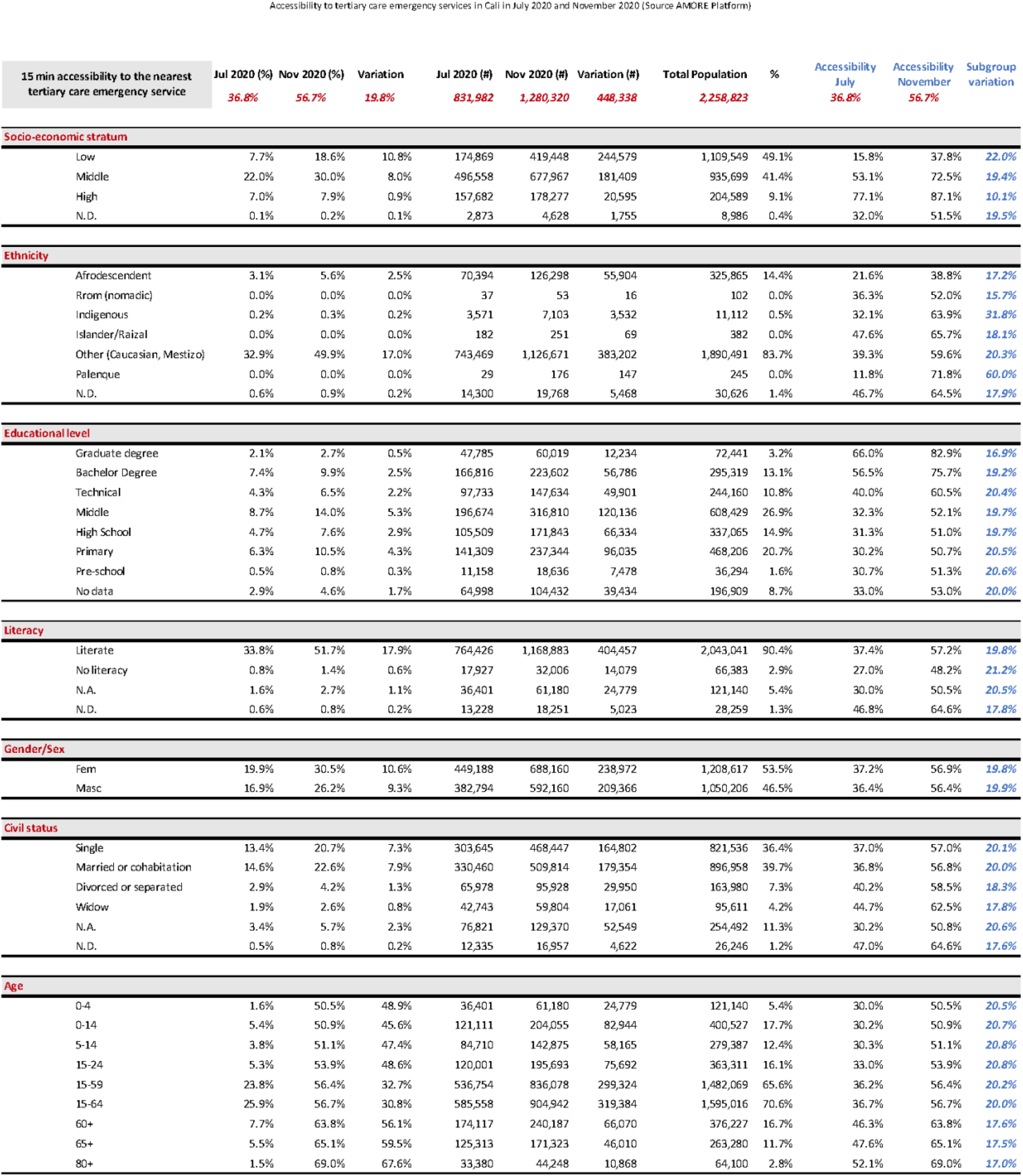
Accessibility to Emergency Care comparing July and November 2020

## Discussion

### Key results

The analysis shows substantial variations in equitable access to tertiary care emergency services due to traffic congestion and the impact that social determinants of health might have on accessibility. The two points of estimate were for early July and late November 2020, and their substantial variations stress the importance of having updatable sources.

The unusually light traffic congestion of November 2020 might have been due to the mobility restrictions associated with the COVID-19 pandemic. Lighter traffic congestion improved accessibility for an additional 448,338 people, most of them living in low-income and low- to mid-income households. These were the people who were also within the 15-minute threshold (Figure 5) and their location can be visualized comparing Map 2 and Map 3. Chart 3 shows accessibility at peak traffic hours disaggregated by sociodemographic characteristics relevant to equity.^35–37^

Easing of traffic congestion brought an additional 22% (244,579) people in low-income household within the 15-minute threshold and 19.4% (181,409) more people living in middle-income households.

Minority ethnic groups like the Palenque Afro-descendants and indigenous people that represent 0.5% of the population of Cali benefitted the most from the traffic congestion reduction. The noticeable improvement among the Palenque resulted from most of their communities being located in the neighborhoods of El Morichal, El Retiro, El Vallado, and Ciudad Córdoba that fell within the 15-min threshold as traffic improved in November Figure 9 Locations of the Palenque people.

**Figure 9.**
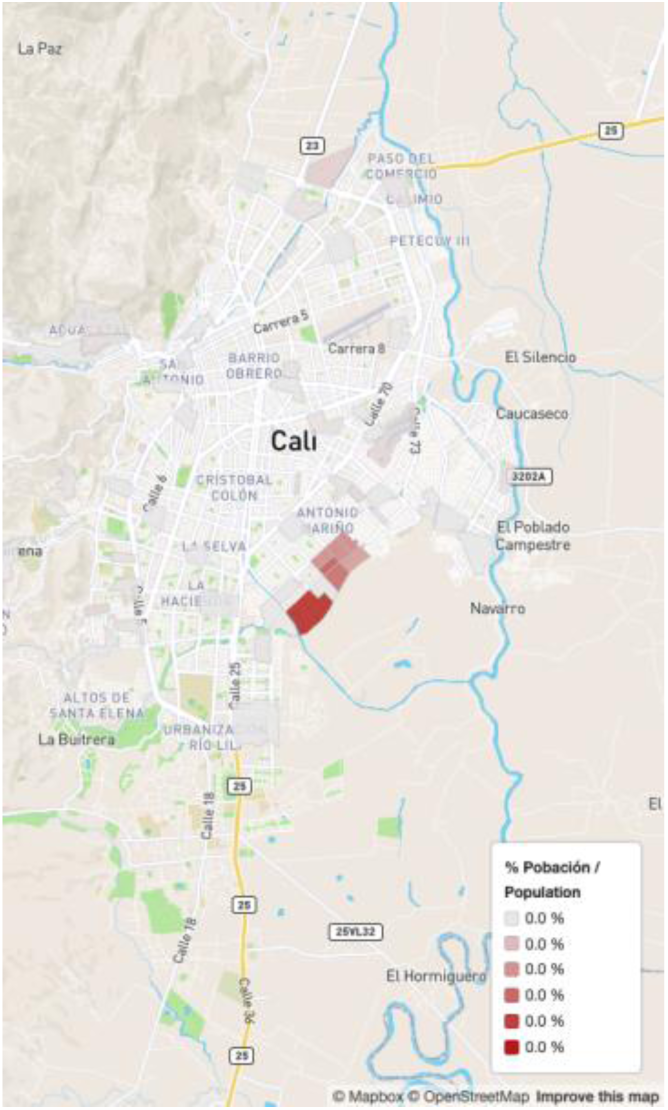
Locations of the Palenque people

Improvements were not significantly disparate among the different groups in terms of sex, educational attainment and literacy, age, and civil status. In terms of education, people with higher educational attainment (a bachelor’s degree or higher) were less impacted by traffic changes (69,020 people benefitted) and those with lower educational attainment were more highly impacted (e.g., 282,505 people with primary, middle, or high school education). Although the variations are not dramatic in relative figures, the absolute numbers are high considering that an additional 332,406 people with primary, middle, high, or technical school education were included as congestion eased.

Comparing age groups, children and the young and working-age populations gained more accessibility with the changes in traffic, as the elderly tend to live closer to health services.

The distance and congestion within that distance impact more on the poor, on people with lower educational attainment, and in the age brackets of children, the youth and working-age population.

Variations in congestion resulted in a substantial improvement, nearly half a million more people being within a 15-minute accessibility threshold, nearly 20% of the population.

Despite improved accessibility, accessibility consistently remained lower for people in low-income households, those without a college education, Afro-descendants and indigenous communities, the young and working-age people, and residents of peripheral areas. Map 2 and Map 3 illustrate that tertiary care health services are far from where most of the population lives. Geospatial analysis, big data, and predictive and prescriptive analytics could be used to inform service planning in ways that maximize accessibility if new services are to address these limitations.

Planners and service providers who want to combat social injustices must examine this new evidence that distance and congestion combine to exclude the most vulnerable and socioeconomically disadvantaged from critical health services. Planners and service providers must then consider bringing services closer to these populations. This new evidence creates an opportunity to make a difference for the future, correct injustices, and help people feel they are included and heard in community planning.

### Limitations

The AMORE Platform uses data modelling and clustering to estimate travel times between the origin and destination sectors, lowering the cost of big data. Operational costs are thus low and platform updates for monitoring and evaluation are affordable. The tradeoff for affordability is that small imprecisions in estimates are possible. These imprecisions are more likely to correspond to populations living near the 15-minute threshold and those further away from TAZ centroids with heavy traffic congestion.

Predicted travel times from the Google Distance Matrix API are known to be accurate and are fed by bigdata from smartphones. Other databases, such as [Waze Transport SDK API](https://developers.google.com/waze/intro-transport) can be used for estimates. Providers do not release prediction algorithms, making it impossible to know the magnitude and variations introduced by unforeseen events, like restrictions associated with the pandemic, and to estimate errors or biases thus introduced on estimates.

Colombian law requires hospitals to treat patients in emergencies. Modeling for this study assumed that people would always go to the nearest hospital in an emergency, but they may not. People may go to a more distant hospital if they know it better, their insurer recommended it, or it has a good reputation. AMORE Platform estimates may thus be more optimistic than reality.

The study assesses travel times from the place of residence to the nearest hospital. Although the maps allow to look at the travel times from a specific sector, the figures do not reflect the fact that the origin of a trip will not always be the place of residence, and that this limitation will spread differently at different moments and for different populations.

### Interpretation

The AMORE Platform reveals accessibility and its health equity implications, providing new dynamic data that accounts for the effects of traffic. It does so with more precision and at a fraction of the costs of household surveys and origin-destination studies, providing a new tool to inform service plans, programs, and policies.

The use of a platform that integrates publicly available data from public sources might be a breakthrough that improves evidence-informed decisions regarding the location and provision of health services. Visualizations might help stakeholders to interpret the data and agree on a common objective and metric: painting the city green by covering its entire population and offering equitable accessibility to all people.

Updating the AMORE Platform is cheaper and faster than updating other origin-destination studies, its assessments are sensitive to variations, and it can be used to monitor evaluate changes. In emergencies such as earthquakes, the platform can show how to optimize the location of emergency services by feeding real-time data downloads rather than predictions.

These findings suggest that with congested traffic in peak hours, most (63%) Cali residents are beyond the 15-minute travel time threshold by car to the nearest tertiary care emergency department. However, this figure fell to 43% when traffic congestion eased.

Reduced accessibility is unevenly distributed and reflects the inverse care law: people who live in low-income households or have less education face longer travel times to tertiary care emergency departments. Incidentally, heavy traffic also affects people on the periphery of Cali, including some high-income households, as congestion clogs roads they use to reach tertiary emergency care facilities.

Accessibility is one of many potential access barriers to health services, and a critical one. Other factors that affect access to health care (e.g., rights, quality, or supplies) are meaningless if patients cannot reach tertiary emergency care in a crisis. Other access barriers to health services (such as non-compliance with Colombian law, quality, and institutional and reputation) are beyond the scope of this study but also merit consideration.

Researchers and planners can use data mining to optimize locations for new tertiary care emergency services and maximize coverage of populations. Data mining can inform construction of new institutions or improvements to existing ones. Optimizing accessibility could inform sound choices. This data is new and provides an opportunity to improve health services planning. Stakeholders and health equity advocates should consider encouraging the integration of accessibility considerations in urban planning processes.

### Generalizability

This study is reproducible in other settings with dynamic travel time data (e.g., from Waze or Google Maps) and georeferenced service and population data that make situational analyses accessible. The accuracy of information depends on the accuracy of its sources (i.e., census data, service providers, travel time estimates or assessments) and the modeling used to make searches and maintenance affordable and data easy to interpret.

## Supporting information

Strobe Checklist

## Data Availability

All data produced in the present study are available upon reasonable request to the authors

https://developers.google.com/maps/documentation/distance-matrix/overview?hl=en_US

https://prestadores.minsalud.gov.co/habilitacion/

https://www.iquartil.net/proyectoAMORE/

https://microdatos.dane.gov.co/index.php/catalog/643

## Other information

### Funding

This study has not yet received external funding; costs have been covered by the principal investigator plus supplementary in-kind contributions by IQuartil SAS, and by Team33 as part of their training with the DS4A data science program. LGC covered the cost of downloading bigdata and the labor costs of producing an advanced prototype of the AMORE Platform. The principal investigator has contributed to this study in his personal capacity and time; his contributions do not necessarily reflect the policies or decisions of his employer.

### Ethical considerations

This observational study integrates anonymized coded secondary data sources obtained from publicly available open records. Colombia’s census microdata used in this study and the location of approved selected health services obtained from REPS are publicly available.^38,39^ This study has not been subject to ethical review.

## Contributions and acknowledgements

### Authors and contributors

The project and manuscript writing were led by Luis Gabriel Cuervo, the principal investigator and project coordinator. Substantive additional contributions and editing of the report were provided by (in alphabetical order): María Olga Bula, Daniel Cuervo, Janet Hatcher-Roberts, Ciro Jaramillo Molina, Eliana Martínez Herrera, Luis Fernando Pinilla, Felipe Piquero, and Lyda Osorio. All members of the AMORE Project Collaboration listed in Table 3 provided comments, conceptual contributions, or consumer perspectives. All those listed approved the manuscript and declared they stood by this research report.

**Table 3.**
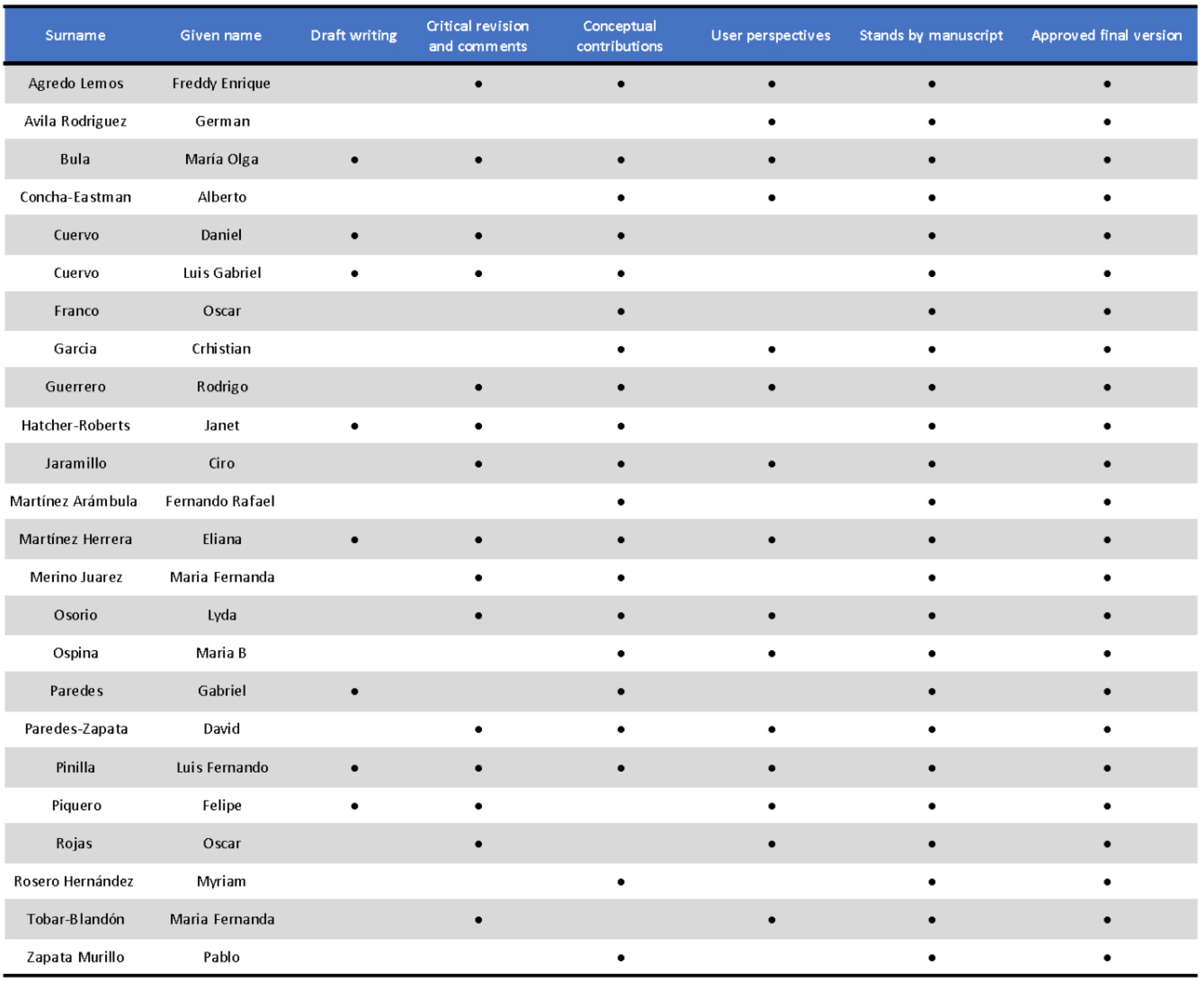
Contributions by Members of the AMORE Project Collaboration approving the manuscript

| Surname | Given name | Draft writing | Critical revision and comments | Conceptual contributions | User perspectives | Stands by manuscript | Approved final version |
| --- | --- | --- | --- | --- | --- | --- | --- |
| Agredo Lemos | Freddy Enrique |  | • | • | • | • | • |
| Avila Rodriguez | German |  |  |  | • | • | • |
| Bula | María Olga | • | • | • | • | • | • |
| Concha-Eastman | Alberto |  |  | • | • | • | • |
| Cuervo | Daniel | • | • | • |  | • | • |
| Cuervo | Luis Gabriel | • | • | • |  | • | • |
| Franco | Oscar |  |  | • |  | • | • |
| Garcia | Christian |  |  | • | • | • | • |
| Guerrero | Rodrigo |  | • | • | • | • | • |
| Hatcher-Roberts | Janet | • | • | • |  | • | • |
| Jaramillo | Ciro |  | • | • | • | • | • |
| Martínez Arámbula | Fernando Rafael |  |  | • |  | • | • |
| Martínez Herrera | Eliana | • | • | • | • | • | • |
| Merino Juarez | Maria Fernanda |  | • | • |  | • | • |
| Osorio | Lyda |  | • | • | • | • | • |
| Ospina | Maria B |  |  | • | • | • | • |
| Paredes | Gabriel | • |  | • |  | • | • |
| Paredes-Zapata | David |  | • | • | • | • | • |
| Pinilla | Luis Fernando | • | • | • | • | • | • |
| Piquero | Felipe | • | • |  | • | • | • |
| Rojas | Oscar |  | • |  | • | • | • |
| Rosero Hernández | Myriam |  |  | • |  | • | • |
| Tobar-Blandón | Maria Fernanda |  | • |  | • | • | • |
| Zapata Murillo | Pablo |  |  | • |  | • | • |

## Acknowledgements

We are grateful to Stephen Volante CT, member of the American Translators Association, for his support with editing and to María Fernanda Merino for providing strategic guidance. IQuartil SAS has hosted the AMORE Platform and we are especially grateful to Pablo Zapata and Lizardo Vanegas for improvements to the AMORE Platform and the AMORE Project website. The development of the AMORE Platform used for this study was led by Luis Gabriel Cuervo, Daniel Cuervo, Luis Fernando Pinilla, and Pablo Zapata. An early prototype of the AMORE Platform was developed under the guidance of the principal investigator (LGC) by Team 33 of the 2020 cohort of the Data Science for All – Correlation Once certificate training program.^40,41^ Members of Team 33 included Daniel Cuervo, Catherine Cabrera, Dario Mogollón, Juan G. Betancourt, Stephanie Rojas, Rafael Ropero, and Santiago Tobar.

## Footnotes

(1) “AMORE” from the anagram of the earlier name of the project “*Análisis espacial con Macrodatos para Orientar los Resultados en pro de la Equidad en salud*” [Spatial analysis with bigdata to guide health equity outcomes].

## Notes

### Competing Interest Statement

All authors have completed the ICMJE uniform disclosure form at www.icmje.org/coi_disclosure.pdf and declare: no financial support from any organisation for the submitted work; IQuartil SAS provided technical support to develop the AMORE Platform and was paid by the principal investigator (LGC) for consulting services; DC is a partner at IQuartil SAS and a sibling to LGC. LFP did consultancy at IQuartil SAS until March 2021. LGC contributed to this work in his personal capacity and time. The views expressed in this article do not necessarily represent the decisions or policies of his employer, PAHO/WHO. Reproductions of this article should not include any suggestion that PAHO/WHO endorsed this research or is endorsing any specific organization, services, or products.

### Clinical Protocols

https://thesiscommons.org/4atqc

### Funding Statement

This study has not received any funding.

### Author Declarations

This observational study integrates anonymized coded secondary data obtained from openly available records listed in the manuscript. Oversight of the project has been provided by the Doctoral Programme on Methodology of Biomedical Research and Public Health at the Department of Paediatrics, Obstetrics & Gynaecology and Preventative Medicine at the Universitat Autonoma de Barcelona. Data sources: (1) Colombia's 2018 Census Microdata downloaded from https://microdatos.dane.gov.co/index.php/catalog/643 (2) Ministry of Health Special Registry of Health Services Providers https://prestadores.minsalud.gov.co/habilitacion/

